# Reproductive lifespan and hormonal therapy in relation to later-life neurovascular health: A population-based study of women in the Gothenburg H70-1944 Birth Cohort

**DOI:** 10.64898/2026.02.04.26345605

**Authors:** Giulia Lorenzon, Gemma García-Lluch, Gillian Coughlan, Shireen Sindi, Silvia Maioli, Konstantinos Poulakis, Jenna Najar, Rosaleena Mohanty, Lina Rydén, Sara Shams, Silke Kern, Rachel Buckley, Eric Westman, Ingmar Skoog, Anna Marseglia

## Abstract

**Background:** Women face greater vulnerability to dementia and Alzheimer’s disease (AD), potentially due to estrogen fluctuations across the lifespan. However, its role in vascular brain health is unclear. We investigated associations between lifelong estrogen exposure—endogenous (reproductive span) and exogenous (oral contraceptives [OC], menopausal hormone therapy [MHT])—and late-life vascular brain injury, AD-related atrophy, and *APOE*-ε4 modification.

**Methods and findings:** We included 352 cognitively unimpaired 70-years-old women from the Gothenburg H70-1944 Birth Cohort with brain MRI and 5-year follow-up. Reproductive lifespan was calculated as age at menopause or oophorectomy minus age at menarche. OC and MHT use were self-reported. Outcomes included cerebral small vessel disease (SVD), AD-related cortical thickness, and white-matter integrity (fractional anisotropy). Linear and multinomial regression and mixed-effects models were adjusted for confounders and stratified by *APOE*-ε4.

Longer reproductive span (OR=0.90 [95%CI 0.83–0.98]) and MHT use (OR=0.43 [95% CI 0.20–0.92]) were linked to lower SVD burden, particularly fewer perivascular spaces and microbleeds. OC and MHT were associated with greater white matter integrity, with additive use throughout life showing the highest fractional anisotropy (OR=0.45 [95% CI 0.12–0.78]). MHT use was associated with greater thickness in areas often affected in AD among *APOE*-ε4 carriers (β=0.38 [95% CI 0.01–0.76]) but not in non-carriers. Longer estrogen exposure was linked to stable cortical thickness and WMH trajectories over time.

**Conclusions:** Extended estrogen exposure throughout life—both endogenous and exogenous—appear to support late-life cerebrovascular health in women, with potential genotype-specific neuroprotective effects. Given the current absence of sex-specific prevention guidelines for cognitive disorders, future research should clarify estrogen’s long-term impact on brain health and cognition to inform personalized medicine.

## INTRODUCTION

Nearly two-thirds of all dementia cases affect women, and the total number of people affected is projected to triple in the coming decades (Nichols et al., 2022). Understanding the biological underpinnings of these sex disparities is essential for developing targeted prevention strategies. Despite the promise of anti-amyloid therapies, they appear to offer limited cognitive benefits for women (Bonkhoff et al., 2025; Dyck et al., 2023), highlighting the need of investigating women-specific risk factors and biological mechanisms.

Women show greater vulnerability to Alzheimer’s disease (AD), with earlier and faster tau accumulation (Coughlan et al., 2025), and faster cognitive decline following amyloid-β accumulation compared to men (Buckley et al., 2018). The mechanisms underlying these differences remain unclear but are likely influenced by the interplay between sex chromosomes, and sex hormones (Coughlan et al., 2025). The female reproductive span—defined as the interval between menarche and menopause—typically ranges from 35 to 38 years (Nabhan et al., 2022; Nichols et al., 2006). Longer estrogen exposure throughout life may provide neuroprotection against dementia, while estrogen dysfunction (e.g., abrupt decline or deficiency due to menopause or other conditions) may accelerate neuropathological changes, increasing dementia risk (Gannon et al., 2023; Mosconi et al., 2021).

Proxies of shorter estrogen exposure across the reproductive lifespan (e.g., late menarche, early menopause) have been linked to increased risks of cognitive decline and dementia (Gong et al., 2022; Han et al., 2023). Menopause has also emerged as a critical window in the aging process, with differences in cognitive outcomes for spontaneous and induced (e.g., surgical) transitions (Dobson et al., 2024). While these findings underscore the importance of endogenous estrogen exposure, the role of exogenous sources remains less clear (Rocca et al., 2024). Some studies reported menopausal hormonal therapy (MHT) associate with reduced dementia risk (Mosconi, Andy, et al., 2025; Mosconi, Nerattini, et al., 2025), whereas others suggest the opposite (Pourhadi et al., 2023). Much of the existing research has focused on MHT, yet fewer studies have explored the impact of oral contraceptives (OC)—which also deliver exogenous estrogen—on brain health (Song et al., 2023). Given that both exogenous estrogen sources may contribute to neuroprotective benefits, it is reasonable to hypothesize that combined OC and MHT use may exert synergistic effects, potentially extending neuroprotective benefits and supporting brain health into later life. However, no studies to date have specifically examined potential additive effects of OC and MHT in relation to markers of late-life vascular and non-vascular brain health.

Beyond its role in neuroprotection, estrogen also supports cerebrovascular health by promoting vasodilation, reducing inflammation, enhancing angiogenesis, and maintaining blood-brain barrier and white matter integrity (Robert, 2023). Thus, its dysfunction may contribute to cerebrovascular damage, increasing the risk of cerebral small vessel disease (SVD), including white matter hyperintensities volume (WMHV), lacunes, microbleeds, and enlarged perivascular spaces (Duering et al., 2023). Recent animal studies suggested that greater cardiometabolic burden – a key SVD risk factor – may drive WMHV burden predominantly in female mice rather than their male counterparts (Abi-Ghanem et al., 2023). However, the long-term impact of endogenous and exogenous estrogen fluctuations on late-life SVD severity remains largely unexplored. Similarly, some evidence suggests that apolipoprotein-ε4 (*APOE*-ε4)—a major genetic risk factor for AD—may modulate estrogen’s neuroprotective effects, with female carriers showing more AD biomarkers burden and mixed responses to hormonal therapies; however, findings are inconsistent (Ambikairajah et al., 2024; Dunk et al., 2025).

In this study, we aimed to examine how both endogenous (reproductive lifespan) and exogenous (OC and MHT) estrogen exposure throughout life are associated with late-life cerebrovascular health and AD-related brain atrophy in a community-based cohort of 70-years-old Swedish women. We further investigated the combined effects of OC and MHT use and whether the presence of *APOE*-ε4 genotype modified such associations.

## METHODS

### 2.1 Study population and setting

Data were drawn from the Gothenburg H70-1944 Birth Cohort, a longitudinal study of 1203 individuals born in 1944 and residing in Gothenborg (Rydberg Sterner et al., 2019). At baseline 2014–2016, participants completed a comprehensive assessment at the Neuropsychiatry Clinic at Sahlgrenska University Hospital or in their homes. These included questionnaires on sociodemographic and lifestyle factors, medical conditions, and women’s reproductive health. Blood and cerebrospinal fluid (CSF) samples were collected and analysed according to standard lab routines at Sahlgrenska University Hospital (Marseglia et al., 2025; Rydberg Sterner et al., 2019). Brain magnetic resonance imaging (MRI) was offered to all participants, with 790 individuals (65.7%) undergoing scanning. For this study, we included 352 women with high-quality FreeSurfer anatomical segmentation of brain MRI, no neurological disorders (i.e., dementia, Parkinson’s disease, multiple sclerosis, epilepsy, brain cancer), and reliable self-reported data on endogenous and exogenous estrogen exposure (**Supplementary Figure 1**). Of these, 286 (81.3%) participated in the follow-up examination at age 75 (2019–2021), of which 205 (58.2%) had available imaging data. The H70 study was approved by the Regional Ethical Review Board in Gothenburg.

### 2.2 Assessment of estrogen exposure

At baseline, trained research staff collected data on women’s reproductive and hormonal history via semi-structured interviews (**Appendix A**), encompassing age at menarche and menopause, menopausal hormonal therapy use and duration, number of pregnancies, and breastfeeding history.

Endogenous and exogenous estrogen exposure variables were derived through a series of steps. First, age at menarche was defined as the age at first menstruation and categorized as early (<12 years), typical (≥12–14 years), or late (>14 years) (Rakic et al., 2024); implausible values (≥20 years) were excluded. Second, age at menopause was defined as 12 months without menstruation and categorized as early (<45 years), typical (≥45–54 years), or late (>54 years) (Jiao et al., 2024); values <38 or >60 years were excluded. Surgical menopause was defined as bilateral oophorectomy—not hysterectomy or unilateral oophorectomy, as these procedures alone do not directly impair ovarian function, where estrogen is primarily produced. To operationalize this, we created a binary variable indicating whether participants had undergone removal of one or both ovaries. Age at surgery was then evaluated in relation to the reported age at last menstruation. If bilateral oophorectomy occurred more than one year prior to menstruation cessation, menopause was considered surgically induced, and age at menopause was replaced with age at surgery to more accurately reflect the onset of estrogen deprivation. Finally, endogenous estrogen exposure throughout life was calculated as reproductive lifespan, that is age at menopause or surgery minus age at menarche (continuous variable).

### 2.3 Brain MRI acquisition and preprocessing

Participants were scanned on a 3.0T Philips Achieva system (Philips Medical Systems) at baseline, and a 3.0T Philips Achieva dStream system (Philips Medical Systems) at the 5-year follow-up. Two human phantoms were scanned before and after the scanner upgrade, with inter-scan intervals of 1.3 and 1.5 years, respectively. The mean percentage change across cortical regions and across both phantoms was −0.67%, which is consistent with previously reported test–retest reliability for structural MRI measures (Treit et al., 2022). Detailed results from FreeSurfer 7.2 for the two phantoms have been previously reported and are provided in Supplementary Table S1 of Lorenzon et al. (2025) (Lorenzon et al., 2025).

The protocol included: a three-dimensional (3D) T1-weighted Turbo Field Echo sequence; a T2-weighted sequence to exclude pathologies like tumors and evaluate enlarged perivascular spaces; a fluid-attenuated inversion recovery (FLAIR) sequence to detect WMHV and lacunes; venous BOLD imaging to detect microbleeds; and a diffusion tensor imaging (DTI) for white matter microstructural integrity. Details on acquisition parameters are detailed elsewhere (Rydberg Sterner et al., 2019). Neuroimaging data were managed and automatically processed through theHiveDB system (Muehlboeck et al., 2014) at Karolinska Institutet. All MRI data underwent quality control by a neuroimaging expert following a standardized protocol (Simmons et al., 2011).

### 2.4 Markers of vascular brain injury and AD-related brain atrophy

SVD markers—including WMHs, lacunes, cerebral microbleeds, and perivascular spaces—were visually rated by a neuroradiologist using standardized scales, following the STRIVE guidelines (Duering et al., 2023). A total SVD score, ranging 0 to 4, was computed by assigning one point for the presence of confluent WMHs (Fazekas’s score 2–3), at least one lacune, at least one cerebral microbleed, and moderate-to-severe perivascular spaces in basal ganglia (≥11-40) (Staals et al., 2014). This composite reflects overall cerebrovascular burden. Due to limited statistical power, scores of 2 (n=24), 3 (n=8), and 4 (n=0) were grouped as ≥2 for the analysis.

WMHV was estimated using the lesion growth algorithm implemented in the Lesion Segmentation Toolbox (LST v3.0.0) in SPM12 (https://www.fil.ion.ucl.ac.uk/spm/), based on probability maps derived from FLAIR intensity distributions, and adjusted for total intracranial volume (Schmidt et al., 2012).

White matter integrity was assessed using DTI-based fractional anisotropy (FA), processed with the FMRIB Diffusion Toolbox in FSL v6.0.7.6 (https://fsl.fmrib.ox.ac.uk/fsl/fslwiki) (Jenkinson et al., 2012). FA values were averaged across the skeletonized white matter mask (Badji et al., 2022).

Brain morphometry was derived from T1-weighted MRI scans processed with FreeSurfer v7.2. Cortical thickness was extracted using the Desikan-Killiany atlas (Desikan et al., 2006). To capture AD-related neurodegeneration, an AD thickness signature was calculated by averaging bilateral thicknesses of entorhinal, inferior temporal, middle temporal, and fusiform regions, normalized by cortical surface area (Marseglia et al., 2025). To facilitate comparability of effect sizes, both AD thickness signature and FA were Z-standardized, with higher values indicating greater integrity. Data for AD thickness signature and WMHV were also available at five-year follow-up.

### 2.5 Covariates and potential confounders

Data on sociodemographic and lifestyle factors, blood/CSF biomarkers, and reproductive history were collected through semi□structured interviews and clinical examinations (Rydberg Sterner et al., 2019). Medical conditions were identified using a combination of self-reported information, clinical measures, medication records, and linkage to the Swedish National Patient Register.

Sociodemographic and lifestyle variables included education (categorized into compulsory primary, secondary, and higher education, based on Swedish classification standards (Marseglia et al., 2025); smoking status (never vs. current/former); at-risk alcohol consumption (defined as >98 g/week per National Institute on Alcohol Abuse and Alcoholism); physical activity (inactive/light vs. active, including regular or intense training); body mass index (BMI), categorized as underweight (<20 kg/m2), normal (20–<25 kg/m2), overweight (25–< 30 kg/m2), or obese (≥30 kg/m2); and waist circumference, categorized as central obesity if ≥80 cm (Ahlner et al., 2023; Marseglia et al., 2025). Global cognition was assessed through the Swedish version of the Mini-Mental State Examination (MMSE) (Rydberg Sterner et al., 2019).

Medical conditions have been described previously (see Supplementary Table 1 in (Marseglia et al., 2025)). Briefly, they included hypertension (≥140/90 mmHg or antihypertensive use), heart disease (presence of myocardial infarction, angina pectoris, heart failure, or atrial fibrillation), diabetes status (normoglycemia vs. prediabetes, or diabetes), stroke/TIA, and depression (none vs. minor/major). Dementia was used as an exclusion criterion and diagnosed according to DSM-III-R criteria, based on neuropsychiatric examinations, informant interviews, and registry data (Rydén et al., 2025).

Exogenous estrogen exposures included self-reported OC use during the reproductive years and MHT during peri/post-menopausal periods. MHT duration was converted from months to years and cleaned to ensure that age at menopause plus MHT duration did not exceed 70 years. The number of pregnancies was calculated as the sum of biological sons and daughters. Implausible breastfeeding durations were excluded.

AD genetic risk was assessed via *APOE* genotyping from blood samples, with participants classified as ε4 carriers or non-carriers (Marseglia et al., 2021). CSF biomarkers were available for a subset of 118 women who underwent lumbar puncture. Analysed markers included: β-amyloid 42 (Aβ_42_; pathological ≤530 pg/mL), phosphorylated tau at threonine 181 (p-tau; ≥80 pg/mL), neurofilament light (NfL), and CSF/serum albumin ratio (pathological ≥10.2) (Marseglia et al., 2025).

### Statistical analysis

Linear regression models (R package stats, version 3.6.2) were employed to estimate mean differences (β-coefficients) and corresponding 95% confidence intervals (CI) in AD thickness signature or FA in relation to the exposure variables. Endogenous exposure was represented by reproductive lifespan modelled as a continuous variable, whereas exogenous exposure included OC and MHT (both never vs. ever used) in separate regression models. To understand if the effect of endogenous exposure was more closely related to age at menarche or menopause (Luders et al., 2025), both variables were included as exposures in the same model (as continuous variables).

Given the categorical nature of the SVD score (0 = reference vs. 1 and ≥2), multinomial logistic regression models were applied to estimate odds ratios (ORs) and 95% CIs for associations between exposure variables and SVD outcome, using the R package nnet (version 7.3-20). Logistic regression models were used also when individual SVD markers were the outcome variables.

To examine whether OC and MHT exert combined effects, we explored their interplay using two complementary approaches. First, we tested a two-way interaction term (OC × MHT) in the regression models to assess potential multiplicative effects on SVD score, FA, and AD thickness signature (separate outcomes). Second, additive effects through a joint exposure analysis, creating a four-level variable: (1) No use (reference), (2) OC only (only OC use and never MHT); (3) MHT only (only MHT use and never OC); and (4) Both OC and MHT. Multinomial regression models were applied. This dual approach allowed us to capture both multiplicative interactions and additive patterns that could reveal cumulative estrogen exposure effects across the life course.

We tested for a multiplicative interaction between *APOE*-ε4 status and estrogen exposure and then stratified the main regression models by *APOE*-ε4 carrier status. CSF biomarker levels were also compared by exposure of interest using chi-square or Wilcoxon rank-sum tests.

To characterize trajectories of AD-related cortical thickness and WMHV over the 5-year follow-up period, linear mixed-effects models were fitted (*lmer* function from R lme4 package) in the subset of women with longitudinal imaging assessment. Each exposure variable was entered in separate models. Fixed effects included the exposures of interest, time of assessment (baseline vs. follow-up), and their interaction term. A random intercept for individuals was included to account for between-subject variability. Models were estimated using restricted maximum likelihood (REML).

All models examining reproductive span included education, number of pregnancies and total months of breastfeeding (both continuous) as covariates, based on prior evidence linking these factors to dementia risk (Yoo et al., 2020). In models assessing exogenous exposure (OC and MHT), education and cardiometabolic conditions—hypertension, heart disease, and diabetes—were included as potential confounders.

A two-sided *p*-value of < 0.05 indicated statistical significance. All statistical analyses were performed with R 2024.04.1.

## 3. RESULTS

Among the 352 septuagenarian women included in the study, 92.2% had attained secondary or higher education. Approximatively one-third were *APOE*-□4 carriers and cognitive functioning was intact (mean MMSE score = 29.2). Cardiometabolic conditions were prevalent, with 69.6% having hypertension, 47.4% pre-/diabetes, and 13.6% heart disease. SVD were absent in 63.1% of participants. SVD was present in 36.9% of the participants. A total of 98 women (27.8%) exhibited one SVD marker, while 32 (9.1%) had two or more (**Supplementary Table 1**). Regarding reproductive history, two-thirds of participants reported typical ages at menarche or menopause, with an average reproductive span of 37.1 ± 4 6 years. During this period, 72.6% reported OC use. In the peri-/post-menopause, 61.2% had used or were currently using MHT (**Supplementary Table 2**). At follow-up, 11 women died before or during the study, 55 dropped out, 72 did not undergo MRI, and 9 failed imaging preprocessing, leaving 205 women with MRI for longitudinal analyses. Participants differed from deceased and dropouts in educational level, *APOE*-□4 genotype, physical inactivity, and mean cortical thickness, while other baseline characteristics were similar (**Supplementary Table 3)**.

### 3.1 Cross-sectional associations between estrogen exposure, SVD, white matter integrity, and AD-related cortical thickness

In multinomial logistic regression, longer reproductive span was associated with lower SVD burden (OR for SVD ≥2: 0.90 [95% CI 0.83–0.98], p=0.012), independent of education, breastfeeding duration and number of pregnancies (**Table 1**). No significant associations were observed with AD thickness signature or FA. To understand if this association was more closely related to age at menarche or menopause, both variables were included as exposures in one model. Later age at menopause was associated with lower SVD burden (OR for SVD score ≥2: 0.90 [95% CI 0.83–0.98], p=0.011), while age at menarche showed no significant association (OR=1.06 [95% CI 0.82–1.38], p=0.634).

**Table 1.** Association between *endogenous* hormonal exposure (reproductive lifespan) and Alzheimer’s disease signature, small vessel disease, and white matter integrity.

|  | AD thickness signature <sup>a</sup> |  |  |  | Small vessel disease score <sup>b</sup> |  |  |  |  |  |  |  | FA average <sup>a</sup> |  |  |  |
| --- | --- | --- | --- | --- | --- | --- | --- | --- | --- | --- | --- | --- | --- | --- | --- | --- |
|  |  |  |  |  | SVD score = 1 |  |  |  | SVD score ≥ 2 |  |  |  |  |  |  |  |
|  | n | β | 95% CI | p | n | OR | 95% CI | p | n | OR | 95% CI | p | n | β | 95% CI | p |
| **Reproductive lifespan** | 351 | -0.005 | (-0.03, 0.02) | 0.709 | 98 | 1.00 | (0.95-1.06) | 0.981 | 32 | 0.90 | (0.83-0.98) | 0.012 | 334 | -0.01 | (-0.03, 0.01) | 0.457 |

Abbreviations: AD, Alzheimer’s disease; FA, fractional anisotropy.
<sup>a</sup> β-coefficients and 95% CI from linear regression models adjusted for educational attainment, number of pregnancies and total months of breastfeeding.
<sup>b</sup> Odds Ratios (OR) and 95% confidence intervals (CI) from multinomial logistic regression adjusted for educational attainment, number of pregnancies and total months of breastfeeding. Reference group was SVD = 0 (n = 221).

Regarding exogenous estrogen exposure, both OC (β = 0.25 [95% CI 0.01–0.50], p=0.039) and MHT use (β = 0.25 [95% CI 0.03–0.47], p=0.024) were associated with FA, independent of education and cardiometabolic conditions (**Table 2**). Only MHT use was associated with lower SVD burden (OR for SVD=1: 0.57 [95% CI 0.35–0.92], p = 0.023; OR for SVD ≥2: 0.43 [95% CI 0.20–0.92], p = 0.029). No associations emerged between OC or MHT use and AD thickness signature.

**Table 2.** Association between *exogenous* hormonal exposure – oral contraceptives and menopausal hormonal therapy – and Alzheimer’s disease signature, small vessel disease, and white matter integrity.

|  | AD thickness signature <sup>a</sup> |  |  |  | Small vessel disease score <sup>b</sup> |  |  |  |  |  |  |  | FA average <sup>a</sup> |  |  |  |
| --- | --- | --- | --- | --- | --- | --- | --- | --- | --- | --- | --- | --- | --- | --- | --- | --- |
|  | n | β | 95% CI | p | SVD score = 1 |  |  |  | SVD score ≥ 2 |  |  |  | n | β | 95% CI | p |
|  |  |  |  |  | n | OR | 95% CI | p | n | OR | 95% CI | p |  |  |  |  |
| OC |  |  |  |  |  |  |  |  |  |  |  |  |  |  |  |  |
| Never | 96 | Reference |  |  | 24 | Reference |  |  | 10 | Reference |  |  | 92 | Reference |  |  |
| User | 256 | -0.06 | (-0.30, 0.18) | 0.633 | 74 | 1.20 | (0.69-2.09) | 0.514 | 22 | 0.85 | (0.38-1.93) | 0.704 | 243 | 0.25 | (0.01, 0.50) | 0.039 |
| MHT |  |  |  |  |  |  |  |  |  |  |  |  |  |  |  |  |
| Never | 136 | Reference |  |  | 46 | Reference |  |  | 17 | Reference |  |  | 132 | Reference |  |  |
| User | 216 | 0.11 | (-0.11, 0.32) | 0.335 | 52 | 0.57 | (0.35-0.92) | 0.023 | 15 | 0.43 | (0.20-0.92) | 0.029 | 203 | 0.25 | (0.03, 0.47) | 0.024 |

Abbreviations: AD, Alzheimer’s disease; FA, fractional anisotropy; MHT, menopausal hormonal therapy; OC, oral contraceptives.
<sup>a</sup> β-coefficients and 95% CI from linear regression models adjusted for educational attainment and cardiometabolic conditions (hypertension, heart disease, diabetes).
<sup>b</sup> Odds Ratios (OR) and 95% CI from multinomial logistic regression adjusted for educational attainment and cardiometabolic conditions (hypertension, heart disease, diabetes). Reference group was SVD = 0.
N for SVD = 0 (reference category) is 73 for MHT non-users, 149 for MHT users, 62 for OC non-users, 160 for OC users.

Analysis of individual SVD markers (WMHs, lacunes, cerebral microbleeds, and perivascular spaces) revealed that MHT use—but not OC—was associated with lower odds of perivascular spaces in basal ganglia (OR 0.57 [95% CI 0.33–1.00], p = 0.048) and cerebral microbleeds (OR=0.45 [95% CI 0.24–0.87], p=0.018) (**Supplementary Table 4**). The association with confluent WMHs (Fazekas’s score 2-3) was borderline (OR=0.55 [95% CI 0.30–1.01], p = 0.055). Later menopause age—but not age at menarche or reproductive span—was associated with reduced odds of lacunes (OR=0.89 [95% CI 0.80–0.98], p=0.023).

### 3.2 Combined effects of OC and MHT on SVD, white matter integrity, and AD-related cortical thickness

For interaction analyses, we dichotomized the SVD score into 0 (no SVD) and ≥1 (at least one SVD) to improve statistical power. In multiplicative interaction models, the OC × MHT term was not significant for any outcome (AD thickness signature: p=0.564; SVD≥1: p=0.389; FA: p=0.982). In joint-exposure analysis, women who used both OC and MHT had greater FA compared with non-users (β=0.45 [95% CI 0.12–0.78], p=0.008). Women who used only MHT had lower odds of SVD≥1 than non-users (OR=0.28 [95% CI 0.10–0.79], p=0.016). A similar trend was observed for combined OC and MHT use, although statistical power was limited. No significant associations were found for AD thickness signature (**Figure 1, Supplementary Table 5**).

**Figure 1.**
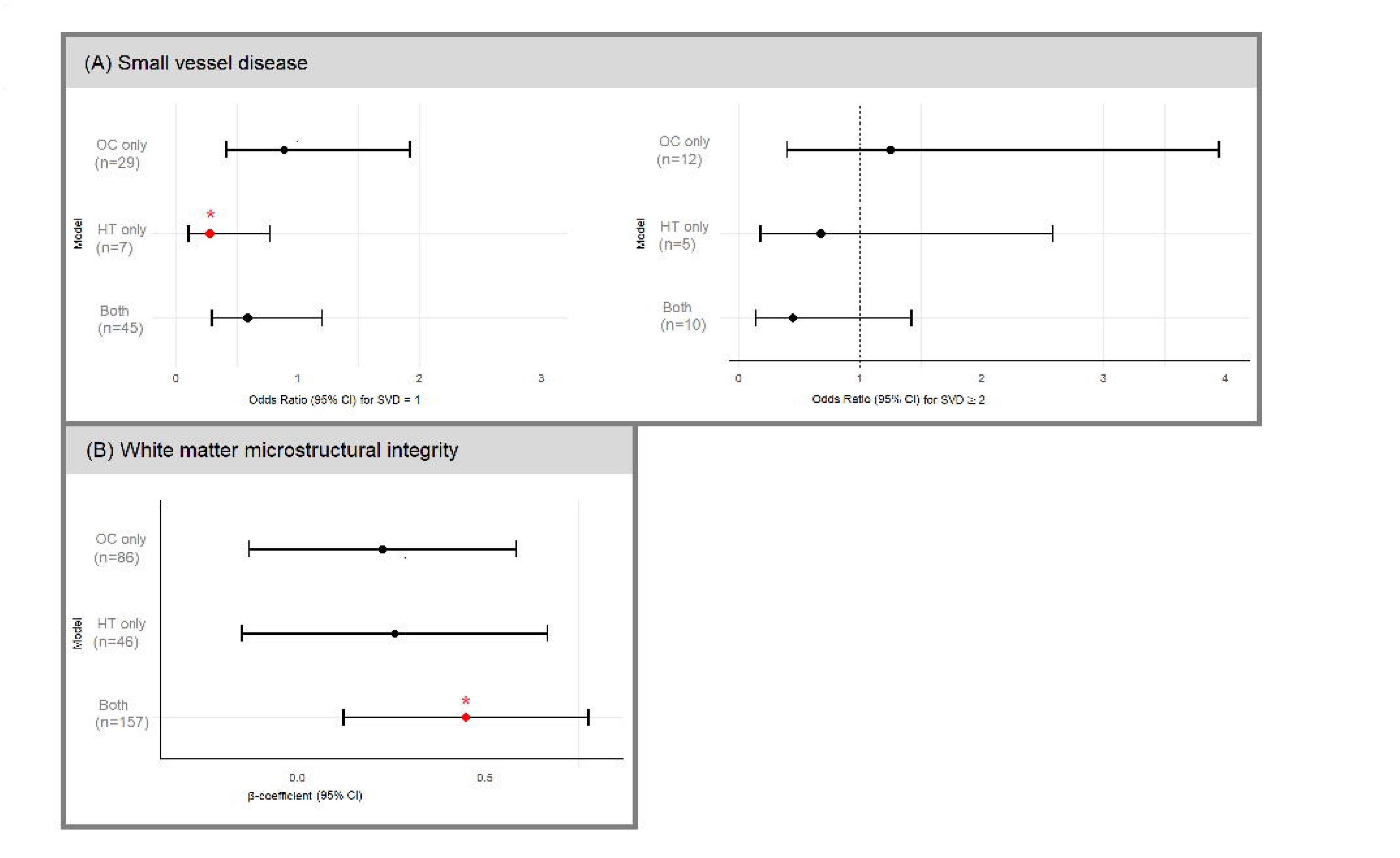
Forest plots depict the joint effect of oral contraceptive (OC) use and menopausal hormonal therapy (MHT) on cerebral small vessel disease (SVD) markers and white matter microstructural integrity (Fractional Anisotropy, FA) in H70 women (n = 352). Multinomial logistic regression models were used for SVD outcomes, presenting odds ratios (ORs) and 95% confidence intervals (CIs). Linear regression models were applied to white matter microstructural integrity, and results are presented as standardized beta coefficients (B) with 95% CIs. The reference group consists of women who reported no use of either OC or MHT (n=17 for SVD=1; n=5 for SVD≥2; n=46 for FA). Stars (*) indicate statistical significance (p > 0.05).

### 3.3 Modifying role of *APOE*-ε4

To assess whether associations between estrogen exposures and neuroimaging markers differed by genetic risk for AD, we took a two-step approach. First, we tested for multiplicate interactions between *APOE*-ε4 carriers’ status and MHT use, which yielded a statistically significant interaction (p = 0.037). Subsequently, we conducted stratified analyses that showed an association between MHT use and greater thickness in AD vulnerable areas exclusively among ε4 carriers (β = 0.38 [95% CI 0.01–0.76], p = 0.045), while no such effect was observed in non-carriers (**Table 3**).

**Table 3.** Stratified analysis by *APOE*-ε4 status in relation to Alzheimer’s disease signature.

| | *APOE*-ε4 non carriers<br>(n=236) | | | *APOE*-ε4 carriers<br>(n=106) | | |
| --- | --- | --- | --- | --- | --- | --- |
| | n | $\beta$ (95% CI) | p | n | $\beta$ (95% CI) | p |
| **Reproductive span** | 236 | -0.001 (-0.03, 0.03) | 0.978 | 106 | -0.003 (-0.05, 0.04) | 0.880 |
| **Oral contraceptive** |  |  |  |  |  |  |
| Never | 68 | Reference |  | 25 | Reference |  |
| User | 168 | -0.002 (-0.29, 0.29) | 0.991 | 81 | -0.03 (-0.48, 0.43) | 0.904 |
| **Menopause**<br>**hormonal therapy** |  |  |  |  |  |  |
| Never | 86 | Reference |  | 47 | Reference |  |
| User | 150 | -0.08 (-0.35, 0.19) | 0.575 | 59 | 0.38 (0.01, 0.76) | 0.045 |

Abbreviations: *APOE*- ε4, apolipoprotein E gene- ε4 allele; MHT, menopausal hormonal therapy; OC, oral contraceptives.
β-coefficients and 95% CI from linear regression models. Models assessing reproductive span were adjusted for education, number of pregnancies, and total months of breastfeeding (continuous). Models examining OC and MHT were adjusted for education and cardiometabolic conditions (hypertension, heart disease, and diabetes).

No differences were observed between *APOE*-ε4 carriers and non-carriers in the association between longer reproductive span and any of the biomarkers. Associations of MHT and OC use with SVD and FA remained directionally consistent but lost statistical significance when stratified, likely due to reduced power (**Supplementary Table 6**).

### 3.4 Cross-sectional associations between estrogen exposure and CSF biomarkers

No differences were observed between estrogen exposures and abnormal AD or neurodegenerative CSF biomarkers (**Supplementary Table 7**).

### 3.5 Five-year longitudinal changes in WMHs volume and AD-related cortical thickness in relation to estrogen exposure

Mixed-effect models showed no longitudinal associations between estrogen exposures and 5-year changes in AD thickness signature and WMHV among 205 women with follow-up MRI data. However, MHT users showed a trend toward slower WMHV burden accumulation (β_slope_ = −1.01, p = 0.087) (**Table 4, Supplementary Figure 2**).

**Table 4.**
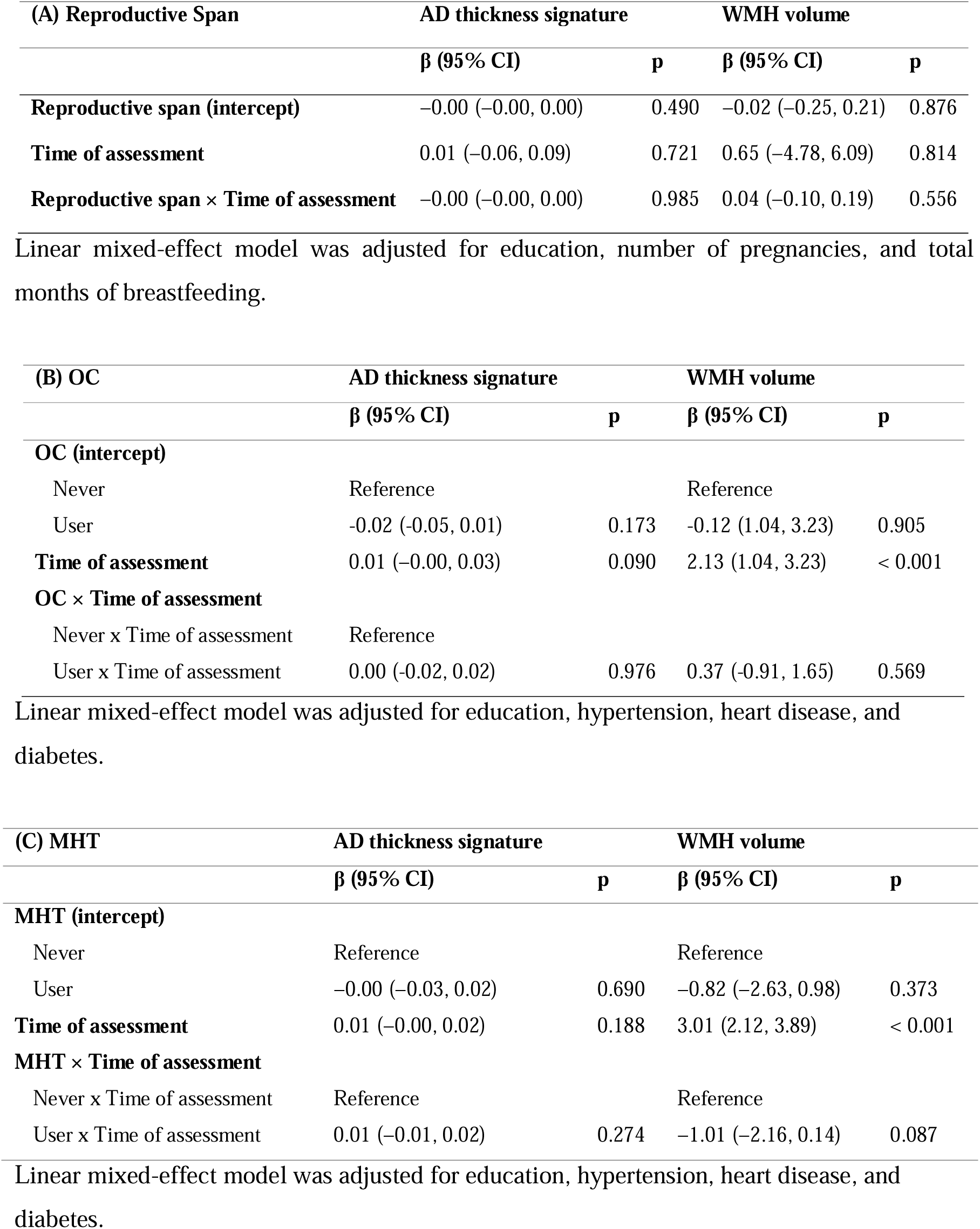
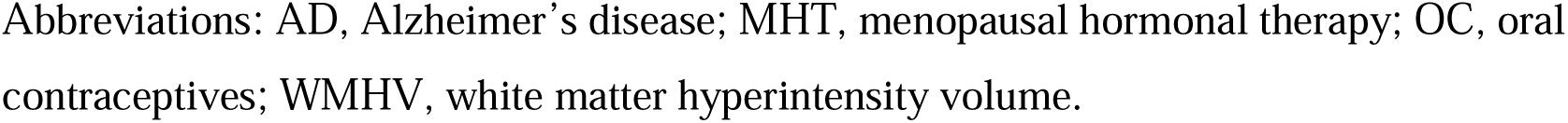
Linear mixed-effects models β-coefficients and 95% confidence intervals (95% CIs) for associations between reproductive span (A), oral contraceptive (B), and menopausal hormonal therapy use (C) with changes in AD thickness signature and WMH volume over five-years follow-up.

## DISCUSSION

In this population-based study of 70-year-old Swedish women, we examined how both endogenous and exogenous estrogen exposure relate to late-life cerebrovascular health and AD-related atrophy. Key findings suggest that longer reproductive span—particularly later menopause—and use of OC and MHT were associated with greater white matter integrity (FA) and/or lower SVD burden, especially fewer perivascular spaces and cerebral microbleeds. Joint exposure to both OC and MHT was linked to greater white matter integrity, possibly reflecting cumulative or sustained estrogen exposure throughout life, though limited by statistical power. No associations were observed with AD-related cortical thickness, except for greater AD-related cortical thickness among ε4 carriers who are using or have used MHT. No association emerged between estrogen exposures and CSF biomarkers.

Our findings indicate that a longer natural exposure to endogenous estrogens—reflected in later menopause or extended reproductive lifespan—is associated with lower burden of overt SVD in late life. Similarly, exogenous estrogen exposure—via OC, MHT, or both—showed comparable associations with cerebrovascular health, as evidenced by greater FA and lower SVD burden, particularly fewer cerebral microbleeds (i.e., small hemorrhages) and perivascular spaces (i.e., fluid-filled compartments surrounding blood vessels that may dilate with aging or pathology). These SVD markers are increasingly recognized not only as contributors to cognitive impairment and dementia (Badji et al., 2023) but also as key components of the brain’s waste clearance systems, including the glymphatic pathway (Solé-Guardia et al., 2025). In our study, the observed lower SVD burden among estrogen-exposed women may reflect estrogen’s role in maintaining vascular integrity and supporting glymphatic function—a hypothesis that warrants further investigation. Previous studies have linked longer reproductive span, earlier menarche, and later menopause to reduced brain aging, reinforcing the neuroprotective role of estradiol (Luders et al., 2025). The distinction between natural versus surgical menopause is also important: while natural menopause involves a gradual decline in estrogen, surgical menopause leads to abrupt hormonal loss, which has been linked to accelerated brain atrophy, potentially related to cardiometabolic dysregulation and inflammation (Gannon et al., 2023; Mosconi et al., 2021). Although reproductive factors such as parity and breastfeeding remain less well understood in relation to late-life brain/cognitive health (Fox et al., 2021; Puri et al., 2024), they were accounted for in our analyses.

While some studies have linked MHT use to accelerated cognitive decline and dementia (Gong et al., 2022; Lee et al., 2024; Rocca et al., 2024), our findings, conversely, offer additional insights into the cerebrovascular pathways potentially influenced by MHT. Using a composite SVD score, we found that MHT use was associated with lower odds of perivascular spaces and microbleeds in late life, suggesting these may be key markers of MHT-related cerebrovascular effects in older women. Contrary to prior studies associating MHT with reduced WMHs (Kantarci et al., 2018), we observed only a borderline association (p=0.055), possibly due to the low prevalence (14%) of confluent WMHs (Fazekas’s score 2-3) in our cohort. Evidence suggests that MHT may exert neuroprotective effects when initiated near menopause (Faubion et al., 2025), with studies reporting cortical preservation and reduced β-amyloid deposition (Mosconi, Nerattini, et al., 2025; Rocca et al., 2024). Our findings extend this perspective by indicating potential cerebrovascular benefits that could mitigate vascular contributions to cognitive impairment. Yet, we were unable to assess whether timing of MHT initiation—closer to menopause or many years later—modulates its effects or interacts with neurodegenerative processes, a hypothesis supported by the notion that estrogen may benefit healthy but not compromised neurons (Brinton, 2008). Future studies need to investigate this “window of opportunity” and the interplay between vascular and neurodegenerative mechanisms during such critical period in modulating MHT effects on brain and cognitive outcomes. Although our analyses were adjusted for education and cardiometabolic comorbidities, associations remained robust, suggesting these do not fully account for the observed effects. Nonetheless, key factors such as MHT formulation, dose, route, duration, and genetic background were unavailable and warrant further investigation.

Our study found that OC use during the reproductive years was associated with greater FA, reflecting preservation of white matter tracts. Compared to MHT, OC use has received far less attention in aging research, and existing findings on its long-term brain effects are inconsistent. While some studies align with our results (Kim et al., 2022; Lee et al., 2024), others report no effects (Faubion et al., 2025) or partial overlap. For example, Haase and colleagues (Haase et al., 2025) found altered white matter connectivity in OC users, with increased connectivity in some networks (e.g., dorsal attention) but also decreased connectivity in others (e.g., somatomotor, visual). Such discrepancies may reflect differences in study design (observation vs. intervention), timing of exposure and outcome assessments, the lag between them, as well as OC formulation (e.g., older vs. modern low-dose ethinyl estradiol), age at initiation (adolescence vs. adulthood), and menstrual cycle regulation (natural vs. hormonal).

Our results also suggest that OC use during the reproductive years and MHT in later life may act cumulatively to support white matter integrity, as reflected in higher FA. This partially supports our initial hypothesis of a cumulative benefit from exogenous estrogen exposure throughout life, whereby early OC use may initiate protective effects on white matter that are sustained or amplified by subsequent MHT. In contrast, interruption of estrogen exposure—such as OC use without later MHT—may limit these long-term benefits. Together, these findings highlight the importance of integrating a life-course perspective on women’s hormonal fluctuations, and the need to assess combined and sequential use of estrogen-based therapies for late-life brain health.

Although no overall association between lifelong estrogen exposure and the AD-related cortical thickness was detected—supporting the idea that hormonal effects in this cohort may act primarily through cerebrovascular pathways—stratified analysis by *APOE*-ε4 genotype revealed greater cortical thickness in AD-vulnerable regions among ε4 carriers with prior/current MHT use. At first glance, this seems counterintuitive given that *APOE*-ε4 is a major non-modifiable risk factor for AD, particularly in women (Gravelsins & Galea, 2025). Several explanations are possible. First, *APOE*-ε4 carriers may be both at higher risk and more responsive to MHT, potentially benefiting from estrogen’s synaptogenic and *APOE*-mediated mechanisms (Mosconi, Andy, et al., 2025). These interpretations align with evidence that *APOE*-ε4 status may modulate MHT’s effects on the brain (Gravelsins & Galea, 2025; Saleh et al., 2023), although findings remain inconsistent, and some studies report opposite associations (Jauregi-Zinkunegi et al., 2025). Second, cognitively unimpaired ε4 carriers may exhibit greater cortical thickness early in life, followed by accelerated atrophy once neurodegeneration begins, consistent with antagonistic pleiotropy (Iacono & Feltis, 2019). Further research is needed to clarify whether these patterns reflect true genotype-specific neurobiological effects or differences in disease stage.

Longitudinally, there were no differences in rates of 5-years change in WMH volume or AD-related cortical thickness. A tendency toward slower WMH lesion accumulation in MHT emerged, suggesting that the neurovascular benefits may persist over time. On another hand, participants who completed follow-up were generally healthier than those who dropped out or died, exhibiting higher education, greater physical activity, and slightly lower smoking prevalence. This ‘healthy survivor’ effect may reduce observable changes over time. Additional factors may also explain the lack of significant findings: the relatively short follow-up period (5 years) may be insufficient to capture meaningful neurodegenerative changes in cognitively unimpaired women; the sample’s narrow age range and overall good health may limit variability; and the limited CSF sample size reduces sensitivity to detect subtle biomarker differences. These findings require replication in larger, more diverse cohorts with longer follow-up.

Strengths of our study include a well-characterized, age-homogeneous, population-based cohort with detailed reproductive and clinical information, combined with longitudinal MRI data. This design enables modeling of retrospectively reported estrogen exposure duration throughout life and objectively measured brain outcomes in late life, while minimizing age-related confounding. Limitations include reliance on self-reported proxies for estrogen exposure, which do not capture individual hormone levels (Mosconi, Nerattini, et al., 2025) and may be subject to recall bias and misclassification, potentially underestimating associations. This risk was mitigated by including only cognitively unimpaired women, as reflected by a mean MMSE score above 29. We also lacked detailed information on MHT initiation and cessation, limiting our ability to examine in-depth the timing hypothesis and the role of MHT characteristics beyond use/non-use (Mosconi, Andy, et al., 2025). The small CSF subsample constrained statistical power, limiting analyses to descriptive comparisons. Similarly, longitudinal, neuroradiologist-assessed markers of SVD, were not yet available for this study, although WMH volume was included as a proxy for SVD in longitudinal analyses. Future studies should also examine the specific roles of uni- or bilateral oophorectomy, with or without hysterectomy, and their long-term associations with MRI measures, as these surgical transitions may substantially alter estrogen exposure trajectories.

The generalizability of our findings extends to women with characteristics similar to those of the 70-year-old participants in this study, although the mechanistic insights may extend to broader populations.

## Conclusions

Our findings suggest that both endogenous and exogenous estrogen exposure during reproductive years and peri-/post-menopause may help preserve cerebrovascular integrity, supporting women’s brain health. Although associations with AD-specific neurodegeneration were limited in this cohort of septuagenarian women—who may have passed the critical window for AD onset—the observed positive influence on white matter integrity and SVD burden highlight vascular processes as potentially key mechanisms. Given the continued absence of sex-specific prevention guidelines for cognitive disorders, understanding estrogen’s long-term impact on brain and cognition remains a clinical and public health priority in the era of personalized medicine.

## Data Availability

Access to this original data is available to the research community upon approval by the cohort PI (Skoog). Applications for accessing these data can be submitted through email to the corresponding authors. Code for data analyses is available on request from the corresponding authors: Anna Marseglia and Giulia Lorenzon.

## ACKNOWLEDGEMENTS

We thank all participants and the staff involved in data collection and management of the Gothenburg H70-1944 Birth Cohort.

## CONTRIBUTORS

**Conceptualization:** A Marseglia, G Lorenzon; **Data curation**: G Lorenzon, G García-Lluch, A Marseglia, Mohanty (DTI processing), S Shams (SVD ratings); **Formal data analysis:** G Lorenzon, A Marseglia, K Poulakis; **Methodology:** A Marseglia, G Lorenzon, J Najar, G Coughlan; **Investigation:** A Marseglia, G Lorenzon, I Skoog (PI of H70), S Kern, E Westman (MRI protocol and processing); **Writing-Original draft:** G Lorenzon, A Marseglia**; Writing-Review & Editing**: All authors; **Supervision:** A Marseglia; **Funding acquisition:** A Marseglia; **Resources:** A Marseglia, E Westman; **Visualization:** G Lorenzon.

All authors approved the final manuscript. Marseglia and Lorenzon had full access to and verified all the data in the study and take the responsibility for the integrity of the data and accuracy of the analyses. All authors had access to all study data and had final responsibility for the decision to submit the manuscript for publication.

## DECLARATION OF INTERESTS

Silke Kern has served at scientific advisory boards, speaker and / or as consultant for Roche, Eli Lilly, Geras Solutions, Optoceutics, Biogen, Eisai, Merry Life, Triolab, Novo Nordisk and Bioarctic, unrelated to present study content. The other authors declare that they have no conflicts of interest.

## FUNDING SOURCES

The H70 study was financed by grants from from the Swedish state under the agreement between the Swedish government and the county councils, the ALF-agreement (ALFGBG-1006423, ALF965812, ALF 716681), the Swedish Research Council (2012-5041, 2015-02830, 2013-8717, 2017-00639, 2019-01096, 2022-00882), Swedish Research Council for Health, Working Life and Wellfare (2013-1202, 2018-00471, AGECAP 2013-2300, 2013-2496, 2018-00471), Konung Gustaf V:s och Drottning Victorias Frimurarestiftelse, Hjärnfonden (FO2014-0207, FO2016-0214, FO2018-0214, FO2019-0163, FO2020-0235, FO2021-0213, FO2024-0341), Alzheimerfonden (AF-554461, AF-647651, AF-743701, AF-844671, AF-930868, AF-940139, AF-968441, AF-980935, AF-1012063, AF-1032314), Eivind och Elsa K:son Sylvans stiftelse, Ann-Louise och Sven-Erik Beiglers stiftelse.

Anna Marseglia received funding from the Swedish Research Council (Vetenskapsrådet, starting grant 2025–02101), the Swedish Research Council for Health, Working Life and Welfare (FORTE; grant 2024–00210), the FORTE research center grant [grant number 2025-02071 - “KI Transdisciplinary Research Center for Personalized Dementia Prevention & Care (TraCeDem)], the Center for Innovative Medicine (CIMED, grant FoUI-988254), the Swedish Dementia Foundation (Demensfonden; grant 4–1759/2024), the Swedish Alzheimer’s Foundation (Alzheimerfonden; grant AF-1032968), the Foundation for Geriatric Diseases at Karolinska Institutet (grants 2025–01993, 2024–02114, 2023–01598, 2022–01268), the Loo och Hans Ostermans stiftelsen (grants 2024–02166, 2023–01645, 2022–01255), the Karolinska Institutet Research Foundation (grant 2024–02580), and Gamla Tjännarinor stiftelse (grants 2025-433, 2024-223, 2023-085).

Gillian Theresa Coughlan is supported by AARF-23-1151259 and K99AG083063. Shireen Sindi is supported by Swedish Research Council (Dnr: 2020-02325), Riksbankens Jubileumsfond (Dnr: P21-0173), Wellcome-Leap CARE, Alzheimerfonden, The Rut and Arvid Wolff Memorial Foundation, The Foundation for Geriatric Diseases at Karolinska Institutet, Forte, and Loo and Hans Osterman Foundation for Medical Research.

Silvia Maioli is supported by the Swedish Research Council (2024-03440), Alzheimerfonden (AF-1011030), King Gustaf V:s and Queen Victoria’s Foundation, Margaretha af Ugglas Foundation, The private initiative "Innovative ways to fight Alzheimeŕs disease - Leif Lundblad Family and others", Wellcome-Leap CARE, Gun och Bertil Stohnes Stiftelse, and Stiftelsen Gamla Tjänarinnor.

Jenna Najar was supported by grants from the Swedish state under the agreement between the Swedish government and the county councils, [the ALF-agreement #1], under Grant [ALFGBG971299, ALFGBG-1007553]; [the Swedish Alzheimer Foundation #2] under Grant [AF967865, AF-993736, AF-1011899]; the [Dementia Foundation #3] under Grant [2021, 2023,2024]; and [Stiftelsen Handlanden Hjalmar Svenssons forskningsfond #4] under Grant [HJSV2021039, HJSV2022059, HJSV2023023].

Silke Kern was financed by grants from the Swedish state under the agreement between the Swedish government and the county councils, the ALF-agreement (ALFGBG-1005471, ALFGBG-965923, ALFGBG-81392, ALF GBG-771071). The Alzheimerfonden (AF-842471, AF-737641, AF-929959, AF-939825). The Swedish Research Council (2019-02075, 2019-02075_15),Stiftelsen Psykiatriska Forskningsfonden, The Swedish Brain Foundation FO2024-0097.

Eric Westman was supported the Swedish Research Council (VR) No. 2016-02282, 2021-01861, the Center for Innovative Medicine (CIMED) No. FoUI-954459, FoUI-975174, the regional agreement on medical training and clinical research (ALF) between Stockholm County Council and Karolinska Institutet No. FoUI-952838, FoUI-954893, The Swedish Brain Foundation (Hjärnfonden) No. FO2022-0084, FO2024-0239, The Swedish Alzheimer’s Foundation (Alzheimerfonden) No. AF-967495, AF-980387, The Swedish Parkinson’s foundation (Parkinsonfonden) No. 1557/24, 1521/23, EU Innovative Health Initiative Joint Undertaking (IHI JU) AD-RIDDLE; King Gustaf V:s and Queen Victorias Foundation, The Swedish Dementia Foundation (Demensfonden), Olle Engkvists Foundation (Olle Engkvists Stiftelse) No. 186-0660, 224-0069 as well as Birgitta and Sten Westerberg for additional financial support.

## Notes

### Author Declarations

The H70 study received ethical approval from the Swedish Ethical Review Authority in Gothenburg (Dnr 869-13; 006-14) and in Uppsala (Dnr 2021-04428). Written informed consent was obtained in agreement with the Helsinki Declaration. All participants provided written informed consent, and in cases where an individual was unable to provide consent, this was obtained from their next-of-kin.

