## Supplementary Materials for "Reproductive lifespan and hormonal therapy in relation to later-life neurovascular health: A population-based study of women in the Gothenburg H70-1944 Birth Cohort"

### **APPENDIX A**

### **Interview protocol for women’s health history in** **the Gothenburg H70-1944 Birth Cohort.**

| **QUESTION** | **ANSWER** | **OPERAZIONALIZATION** |
| --- | --- | --- |
| **Age at menarche** | | |
| 1. How old were you when you had your first period? | age | Early menarche: <12 years  Typical menarche: ≥12-14 years  Late menarche: >14 years |
| **Age at menopause** | | |
| 2. How old were you when you had your last period? | age | Early menopause: <45 years  Typical menopause: ≥45-54 years  Late menopause: >54 years |
| 3. Have your ovaries or uterus been surgically removed? | 0=No surgery  1=Ovaries removal  2=Uterus removal  3=Both ovaries and uterus removal  4=One ovary removal  5=One ovary and uterus removal | **Surgical menopause:**  No = No surgery  Yes = Bilateral oophorectomy (answers 1 or 3) |
| 4. At which age did this happen? | age | Age at surgery (continuous) |
| **Oral contraceptive (OC)** | | |
| 5. Have you used oral contraceptive pills? | 0 = Never  1 = Yes | **OC use:**  0 = Never  1 = Prior use |
| 6. At which age did you start? | age | **Duration of OC (years):**  Age at stop – age at start (continuous) |
| 7. At which age did you stop? | age |  |
| **Menopause Hormonal Therapy (MHT)** | | |
| 8. Have you used estrogen medication for symptoms associated with menopause? | 0 = Never  1 = Yes, in the past  2 = Yes, currently | **Menopause MHT use**:  0 = Never  1 = Prior or current use |
| 9. Have you used estrogen medication for symptoms associated with menopause: How many months in total? | Number of months | **Duration of** **Menopause MHT (years):**  Converted in years, and dichotomized in < 5 vs. ≥ 5 years |
| **Other reproductive health information** | | |
| 10. How many months have you breastfed your children in total? | Number of months | **Total months of breastfeeding** (continuous) |
| 11. How many biological daughters/sons do you have? | Number of daughters or sons alive or deceased | **Number of pregnancies**:  Sum of number of daughters and sons (continuous) |

### **Supplementary Table 1.** Baseline characteristics of 352 dementia-free women from the Gothenburg H70-Birth Cohort 1944.

| **Baseline characteristics** | **Mean ± SD or number (%)** |
| --- | --- |
| **Socio-demographic and lifestyle** |  |
| Education |  |
| Primary school | 25 (7.1%) |
| Secondary school | 195 (55.4%) |
| Higher education | 132 (37.5%) |
| *APOE*-ɛ4 carrier | 106 (31.0%) |
| MMSE score | 29.2 ± 1.1 |
| Smoking (current/former) | 209 (59.4%) |
| Alcohol risk consumption | 78 (22.2%) |
| Physical inactivity | 9 (2.6%) |
| BMI, kg/m^2^ |  |
| Underweight (<20) | 24 (6.8%) |
| Normal (≥20–25) | 158 (44.9%) |
| Overweight (≥25–30) | 115 (32.7%) |
| Obese (≥30) | 54 (15.3%) |
| **Cardiometabolic and medical conditions** |  |
| Hypertension (≥140/90 mmHg) | 245 (69.6%) |
| Heart disease | 48 (13.6%) |
| Stroke/TIA | 31 (8.8%) |
| Diabetes status |  |
| Normoglycemia | 185 (52.6%) |
| Prediabetes | 132 (37.5%) |
| Diabetes | 35 (9.9%) |
| Depression (major/minor) | 31 (8.8%) |
| **Neuroimaging** |  |
| Mean cortical thickness (mm) | 2.4 ± 0.1 |
| AD thickness signature (mm) | 2.7 ± 0.1 |
| Lacunes | 18 (5.1%) |
| Confluent WMH | 49 (13.9%) |
| Cerebral microbleeds | 42 (11.9%) |
| Enlarged PVS-Basal ganglia | 62 (17.6%) |
| SVD score |  |
| 0 | 222 (63.1%) |
| 1 | 98 (27.8%) |
| ≥2 | 32 (9.1%) |
| DTI’s average FA | 0.36 ± 0.05 |

Abbreviations: AD, Alzheimer’s disease; *APOE*-ɛ4, apolipoprotein E gene-ɛ4 allele; BMI, body mass index; DTI, diffusion tensor imaging; FA, fractional anisotropy; MMSE, Mini-Mental State Examination; PVS, perivascular spaces; SVD, small vessels disease; TIA, transient ischemic attack; WMH, white matter hyperintensity.

Missing data: MMSE (n=1), Physical inactivity (n=10), BMI (n=1), FA (n=17), Lacunes (n=2), Confluent WMH (n=3), Enlarged PVS-Basal ganglia (n=3), Cerebral microbleeds (n=4).

### **Supplementary Table 2.** Lifetime reproductive factors of 352 dementia-free women from the Gothenburg H70– Birth Cohort 1944.

|  | **Mean ± SD or n (%)** |
| --- | --- |
| Reproductive span | 37.1 ± 4.6 |
| Age at menarche | 13.3 ± 1.5 |
| Early (<12 years) | 43 (12.3%) |
| Typical (≥12-14 years) | 230 (65.5%) |
| Late (>14 years) | 78 (22.2%) |
| Age at natural menopause (considering surgery) | 50.4 ± 4.5 |
| Early (<45 years) | 32 (9.1%) |
| Regular (≥45-54 years) | 240 (68.4%) |
| Late (>54 years) | 79 (22.5%) |
| Number of pregnancies | 1.9 ± 1.1 |
| Months of breastfeeding | 9.5 ± 10.0 |
| Bilateral oophorectomy | 28 (8.0%) |
| **Hormonal Therapies** | |
| **During reproductive lifespan** | |
| OC user (ever) | 255 (72.6%) |
| OC duration, years | 6.7 ± 7.8 |
| **Peri-/post-menopause** |  |
| Menopausal MHT |  |
| Never | 136 (38.7%) |
| Prior | 185 (52.7%) |
| Current | 30 (8.5%) |
| MHT duration |  |
| <5 years | 171 (57.4%) |
| ≥5 years | 127 (42.6%) |

Data are presented as Mean ± Standard deviations or number (proportion %).

Abbreviations: MHT, menopause hormonal therapy; OC, oral contraceptive.

Missing data: Reproductive span (n=1), Age at menopause incl. surgery (n=1), Number of pregnancies (n=7), Months of breastfeeding (n=15), OC duration (n=28), MHT duration (n=54).

### **Supplementary Table 3.** Baseline characteristics of 352 dementia-free women with follow-up data from the Gothenburg H70 Birth Cohort 1944 study, comparing those who completed the 5-year follow-up to those who died or dropped out.

| **Baseline characteristics** | **Alive (n=286)** | **Deceased (n=11)** | **Dropout (n=55)** | **Total (n=352)** | **p** |
| --- | --- | --- | --- | --- | --- |
| **Socio-demographic and lifestyle** | | | | | |
| Education |  |  |  |  | **0.019** |
| Primary school | 16 (5.6%) | 1 (9.1%) | 8 (14.5%) | 25 (7.1%) |  |
| Secondary school | 153 (53.5%) | 6 (54.5%) | 36 (65.5%) | 195 (55.4%) |  |
| Higher education | 117 (40.9%) | 4 (36.4%) | 11 (20.0%) | 132 (37.5%) |  |
| *APOE*-ɛ4 carrier | 79 (28.2%) | 2 (18.2%) | 25 (49.0%) | 106 (31.0%) | **0.008** |
| MMSE score | 29.2 ± 1.0 | 29.2 ± 1.3 | 29.0 ± 1.2 | 29.2 ± 1.1 | 0.356 |
| Smoking (current/former) | 162 (56.6%) | 9 (81.8%) | 38 (69.1%) | 209 (59.4%) | 0.069 |
| Alcohol risk consumption | 66 (23.1%) | 1 (9.1%) | 11 (20.0%) | 78 (22.2%) | 0.502 |
| Physical inactivity | 3 (1.0%) | 1 (9.1%) | 5 (9.1%) | 9 (2.6%) | **0.003** |
| BMI, kg/m^2^ |  |  |  |  | 0.948 |
| Underweight (<20) | 19 (6.6%) | 1 (9.1%) | 4 (7.3%) | 24 (6.8%) |  |
| Normal (≥20–25) | 131 (45.8%) | 5 (45.5%) | 22 (40.0%) | 158 (44.9%) |  |
| Overweight (≥25–30) | 95 (33.2%) | 3 (27.3%) | 17 (30.9%) | 115 (32.7%) |  |
| Obese (≥30) | 40 (14.0%) | 2 (18.2%) | 12 (21.8%) | 54 (15.3%) |  |
| **Cardiometabolic and medical conditions** | | | | | |
| Hypertension (≥140/90 mmHg) | 196 (68.5%) | 8 (72.7%) | 41 (74.5%) | 245 (69.6%) | 0.657 |
| Heart disease | 38 (13.3%) | 4 (36.4%) | 6 (10.9%) | 48 (13.6%) | 0.074 |
| Stroke/TIA | 22 (7.7%) | 2 (18.2%) | 7 (12.7%) | 31 (8.8%) | 0.259 |
| Diabetes status |  |  |  |  | 0.142 |
| Normoglycemia | 157 (54.9%) | 3 (27.3%) | 25 (45.5%) | 185 (52.6%) |  |
| Prediabetes | 104 (36.4%) | 5 (45.5%) | 23 (41.8%) | 132 (37.5%) |  |
| Diabetes | 25 (8.7%) | 3 (27.3%) | 7 (12.7%) | 35 (9.9%) |  |
| Depression (major/minor) | 24 (8.4%) | 1 (9.1%) | 6 (10.9%) | 31 (8.8%) | 0.833 |
| **Neuroimaging** |  |  |  |  |  |
| Mean cortical thickness (mm) | 2.4 ± 0.1 | 2.3 ± 0.1 | 2.4 ± 0.1 | 2.4 ± 0.1 | **0.044** |
| AD thickness signature (mm) | 2.7 ± 0.1 | 2.7 ± 0.1 | 2.7 ± 0.1 | 2.7 ± 0.1 | 0.200 |
| Lacunes | 15 (5.2%) | 0 (0.0%) | 3 (5.5%) | 18 (5.1%) | 0.653 |
| Confluent WMH | 39 (13.6%) | 3 (27.3%) | 7 (12.7%) | 49 (13.9%) | 0.651 |
| Cerebral microbleeds | 32 (11.2%) | 1 (9.1%) | 9 (16.4%) | 42 (11.9%) | 0.793 |
| Enlarged PVS-Basal ganglia | 49 (17.1%) | 1 (9.1%) | 12 (21.8%) | 62 (17.6%) | 0.712 |
| SVD score |  |  |  |  | 0.922 |
| 0 | 184 (64.3%) | 7 (63.6%) | 31 (56.4%) | 222 (63.1%) |  |
| 1 | 77 (26.9%) | 3 (27.3%) | 18 (32.7%) | 98 (27.8%) |  |
| ≥2 | 25 (8.7%) | 1 (9.1%) | 6 (10.9%) | 32 (9.1%) |  |
| DTI’s Average FA | 0.4 ± 0.0 | 0.3 ± 0.0 | 0.3 ± 0.0 | 0.4 ± 0.0 | 0.131 |

Data are presented as mean ± standard deviations (SD) or number (proportion %).

Abbreviations: AD, Alzheimer’s disease; *APOE*-ɛ4, apolipoprotein E gene-ɛ4 allele; BMI, body mass index; DTI, diffusion tensor imaging; FA, fractional anisotropy; MMSE, Mini-Mental State Examination; PVS, perivascular spaces; SVD, small vessels disease; TIA, transient ischemic attack; WMH, white matter hyperintensity.

Missing data: MMSE (n=1), Physical inactivity (n=10), BMI (n=1), FA (n=17), Lacunes (n=2), Confluent WMH (n=3), Enlarged PVS-Basal ganglia (n=3), Cerebral microbleeds (n=4).

### **Supplementary Table 4.** Association between reproductive span, menopausal hormonal therapy, and oral contraceptives use, with MRI markers of cerebral small vessel disease.

|  | **Lacunes** | | |  | **Confluent WMH** | | |  | **Enlarged PVS-BG** | | |  | **Cerebral microbleeds** | | |  |
| --- | --- | --- | --- | --- | --- | --- | --- | --- | --- | --- | --- | --- | --- | --- | --- | --- |
|  | **n** | **OR** | **95% CI** | **p** | **n** | **OR** | **95% CI** | **p** | **n** | **OR** | **95% CI** | **p** | **n** | **OR** | **95% CI** | **p** |
| **Reproductive span (years)** | 349 | 0.91 | (0.82-1.01) | 0.064 | 348 | 0.97 | (0.90-1.04) | 0.341 | 348 | 0.95 | (0.90-1.01) | 0.129 | 347 | 0.97 | (0.90-1.05) | 0.443 |
| **Oral Contraceptive** | | | | |  | | | |  | | | |  | | | |
| Never | 94 | Reference | | | 94 | Reference | | | 94 | Reference | | | 94 | Reference | | |
| User | 256 | 1.23 | (0.39-3.95) | 0.707 | 255 | 1.12 | (0.55-2.28) | 0.759 | 255 | 0.98 | (0.52-1.83) | 0.952 | 254 | 0.65 | (0.33-1.30) | 0.223 |
| **MHT** | | | | |  | | | |  | | | |  | | | |
| Never | 134 | Reference | | | 134 | Reference | | | 134 | Reference | | | 133 | Reference | | |
| User | 216 | 1.27 | (0.46-3.52) | 0.639 | 215 | 0.55 | (0.30-1.01) | 0.055 | 215 | 0.57 | (0.33-1.00) | 0.048 | 215 | 0.45 | (0.24-0.87) | 0.018 |

Data are presented as odds ratios (ORs) and 95% confidence intervals (CIs) from multinomial logistic regression models adjusted for educational attainment and cardiometabolic conditions (hypertension, heart disease, diabetes). Reference group was SVD = 0.

Abbreviations: BG, basal ganglia; MHT, menopausal hormonal therapy; MRI, magnetic resonance imaging; PVS, perivascular spaces; WMH, white matter hyperintensity.

### **Supplementary Table 5.** Joint effect analysis of the effect of oral contraceptives plus menopausal hormonal therapy on small vessel disease and white matter microstructural integrity.

|  |  |  | **Small vessel disease score (SVD)** ^b^ | | | | | | | | **FA average** ^a^ | | | |
| --- | --- | --- | --- | --- | --- | --- | --- | --- | --- | --- | --- | --- | --- | --- |
|  |  |  | **SVD score = 1** | | | | **SVD score ≥ 2** | | | |  | | | |
| **OC** | **MHT** |  | **n** | **OR** | **95% CI** | **p** | **n** | **OR** | **95% CI** | **p** | **n** | **β** | **95% CI** | **p** |
| No | No |  | 17 | Reference | | | 5 | Reference | | | 46 | Reference | | |
| Yes | No |  | 29 | 0.89 | (0.41-1.95) | 0.779 | 12 | 1.24 | (0.39-3.98) | 0.719 | 86 | 0.22 | (-0.13-0.58) | 0.218 |
| No | Yes |  | 7 | 0.28 | (0.10-0.79) | 0.016 | 5 | 0.66 | (0.17-2.60) | 0.556 | 46 | 0.23 | (-0.18-0.64) | 0.271 |
| Yes | Yes |  | 45 | 0.61 | (0.30-1.24) | 0.172 | 10 | 0.44 | (0.14-1.43) | 0.173 | 157 | 0.45 | (0.12-0.78) | 0.008 |

Abbreviations: FA, fractional anisotropy; MHT, menopause hormonal therapy; OC, oral contraceptives; OR, odds ratio; SVD, small vessel disease.

^a^ β-coefficients and 95% CI from linear regression models adjusted for educational attainment and cardiometabolic conditions (hypertension, heart disease, diabetes).

^b^ OR and 95% CI from multinomial logistic regression adjusted for educational attainment and cardiometabolic conditions (hypertension, heart disease, diabetes). Reference group was SVD = 0 (n = 25 none; n = 48 only OC; n = 37 only MHT; n = 112 both).

### **Supplementary Table 6.** Associations of reproductive span (A), oral contraceptives (B) and menopausal hormonal therapy (C) with small vessel disease and fractional anisotropy in *APOE*-ε4 carriers vs. non-carriers.

| **A) CARRIERS (n=139)** | | | |
| --- | --- | --- | --- |
|  | **Small vessel disease score (SVD)** ^b^ | | **FA average** ^a^ |
|  | **SVD score = 1** | **SVD score ≥ 2** | |

|  | **n** | **OR** | **95% CI** | **p** | **n** | **OR** | **95% CI** | **p** | **n** | **β** | **95% CI** | **p** |
| --- | --- | --- | --- | --- | --- | --- | --- | --- | --- | --- | --- | --- |
| **Reproductive Span** | 36 | 0.97 | (0.87–1.07) | 0.502 | 8 | 0.84 | (0.71–1.00) | 0.053 | 101 | -0.003 | (-0.03, 0.02) | 0.842 |

**NON-CARRIERS (n=269)**

|  | **n** | **OR** | **95% CI** | **p** | **n** | **OR** | **95% CI** | **p** | **n** | **β** | **95% CI** | **p** |
| --- | --- | --- | --- | --- | --- | --- | --- | --- | --- | --- | --- | --- |
| **Reproductive Span** | 60 | 1.01 | (0.94–1.08) | 0.875 | 23 | 0.89 | (0.81–0.99) | 0.025 | 224 | -0.01 | (-0.03, 0.02) | 0.580 |

*Note.* n for SVD = 0 (reference category) is 62 for carriers and 152 for non-carriers.

| **B) CARRIERS (n=139)** | | | | | | | | | | | | | | |
| --- | --- | --- | --- | --- | --- | --- | --- | --- | --- | --- | --- | --- | --- | --- |
|  | **Small vessel disease score (SVD)** ^b^ | | | | | | | | | | | **FA average** ^a^ | |  |
|  | **SVD score = 1** | | | | | **SVD score ≥ 2** | | | | | |  | |  |
|  | **n** | **OR** | **95% CI** | **p** | | **n** | **OR** | | **95% CI** | **p** | **n** | **β** | **95% CI** | **p** |
| OC |  |  | | | |  |  | | | |  |  | | |
| Never | 7 | Reference | | | | 2 | Reference | | | | 23 | Reference | | |
| OC user | 29 | 1.26 | (0.44–3.62) | 0.668 | | 6 | 0.77 | | (0.12–4.78) | 0.775 | 78 | 0.41 | (-0.25, 1.06) | 0.222 |
| **NON-CARRIERS (n=269)** | | | | | | | | | | | | | | |
|  | **n** | **OR** | **95% CI** | **p** | **n** | | | **OR** | **95% CI** | **p** | **n** | **β** | **95% CI** | **p** |
| OC |  |  |  |  |  | | |  |  |  |  |  |  |  |
| Never | 17 | Reference | | | | 8 | | Reference | | | 66 | Reference | | |
| OC user | 43 | 1.11 | (0.56–2.19) | 0.768 | 15 | | | 0.82 | (0.32–2.14) | 0.686 | 159 | 0.21 | (-0.03, 0.46) | 0.088 |

*Note.* In carriers, n for SVD = 0 (reference category) is 16 for OC non-users and 46 for OC users. In non-carriers, n for SVD = 0 (reference category) is 43 for OC non-users and 110 for OC users.

| **C) CARRIERS (n=139)** | | | | | | | | | | | | | |
| --- | --- | --- | --- | --- | --- | --- | --- | --- | --- | --- | --- | --- | --- |
|  | **Small vessel disease score (SVD)** ^b^ | | | | | | | | | | **FA average** ^a^ | |  |
|  | **SVD score = 1** | | | | **SVD score ≥ 2** | | | | | |  | |  |
|  | **n** | **OR** | **95% CI** | **p** | **n** | **OR** | | **95% CI** | **p** | **n** | **β** | **95% CI** | **p** |
| MHT |  |  | | |  |  | | | |  |  | | |
| Never | 19 | Reference | | | 4 | Reference | | | | 45 | Reference | | |
| MHT user | 17 | 0.56 | (0.23-1.32) | 0.182 | 4 | 0.77 | | (0.15-3.84) | 0.747 | 56 | 0.38 | (-0.17, 0.92) | 0.175 |
| **NON-CARRIERS (n=269)** | | | | | | | | | | | | | |
|  | **n** | **OR** | **95% CI** | **p** | **n** | | **OR** | **95% CI** | **p** | **n** | **β** | **95% CI** | **p** |
| MHT |  |  |  |  |  | |  |  |  |  |  |  |  |
| Never | 26 | Reference | | | 12 | | Reference | | | 84 | Reference | | |
| MHT user | 34 | 0.58 | (0.31-1-09) | 0.092 | 11 | | 0.41 | (0.16-1.01) | 0.051 | 141 | 0.20 | (-0.02, 0.43) | 0.079 |

*Note.* In carriers, n for SVD = 0 (reference category) is 24 for MHT non-users and 38 for MHT users. In non-carriers, n for SVD = 0 (reference category) is 48 for MHT non-users and 105 for MHT users.

Abbreviations: AD, Alzheimer’s disease; *APOE*-ɛ4, apolipoprotein E gene-ɛ4 allele; FA, fractional anisotropy; MHT, menopausal hormonal therapy; OC, oral contraceptives; SVD, small vessel disease.

^a^ β-coefficients and 95% CI from linear regression models adjusted for educational attainment and cardiometabolic conditions (hypertension, heart disease, diabetes).

^b^ Odds Ratios (OR) and 95% CI from multinomial logistic regression adjusted for educational attainment and cardiometabolic conditions (hypertension, heart disease, diabetes). Reference group was SVD = 0.

### **Supplementary Table 7.** Distribution of abnormal AD/neurodegenerative biomarkers for 118 women with available CSF data.

| **Biomarker** | **Measure** | **Total**  **(n=118)** | **Oral contraceptive** | | | **Menopausal Hormonal Therapy** | | |
| --- | --- | --- | --- | --- | --- | --- | --- | --- |
|  |  |  | **Non users (n=26)** | **Users (n=92)** | ***p*-value** | **Non users (n=39)** | **Users (n=79)** | ***p*-value** |
| **Aβ_42_ (A)** ^a^ | **Abnormal** ≤530 pg/mL | 51 (44.0%) | 10 (38.5%) | 41 (45.6%) | 0.521 | 21 (53.8%) | 30 (39.0%) | 0.127 |
| **p-tau 181 (T)** ^a^ | **Abnormal** ≥80 pg/mL | 4 (3.4%) | 1 (3.8%) | 3 (3.3%) | 0.884 | 1 (2.6%) | 3 (3.8%) | 0.728 |
| **NfL (N)** ^b^ | **Median [IQR]** | 716.5 [314.3] | 623 [211.8] | 771.5 [338.5] | 0.174 | 628 [292.5] | 774 [327] | 0.825 |
| **CSF/Serum Albumin (V)** ^a^ | **Abnormal** ≥10.2 | 4 (3.4%) | 0 (0.0%) | 4 (4.3%) | 0.279 | 1 (2.6%) | 3 (3.8%) | 0.728 |

Abbreviations: Aβ_42_, amyloid-β 42; CSF, cerebrospinal fluid; MHT, menopausal hormonal therapy; NfL; neurofilament light; OC, oral contraceptives; p-tau 181, phosphorylated tau at threonine 181.

Missing data: Aβ_42_ (n = 2); NfL (n = 2).

^a^ Categorical variables are reported as number and percentage [n (%)] from chi-square tests.

^b^ Continuous variables are presented as median and interquartile range [median (IQR)] from Wilcoxon rank-sum tests.

### **Supplementary Figure 1.** Flowchart of study participants in the H70-1944 Birth Cohort brain MRI.

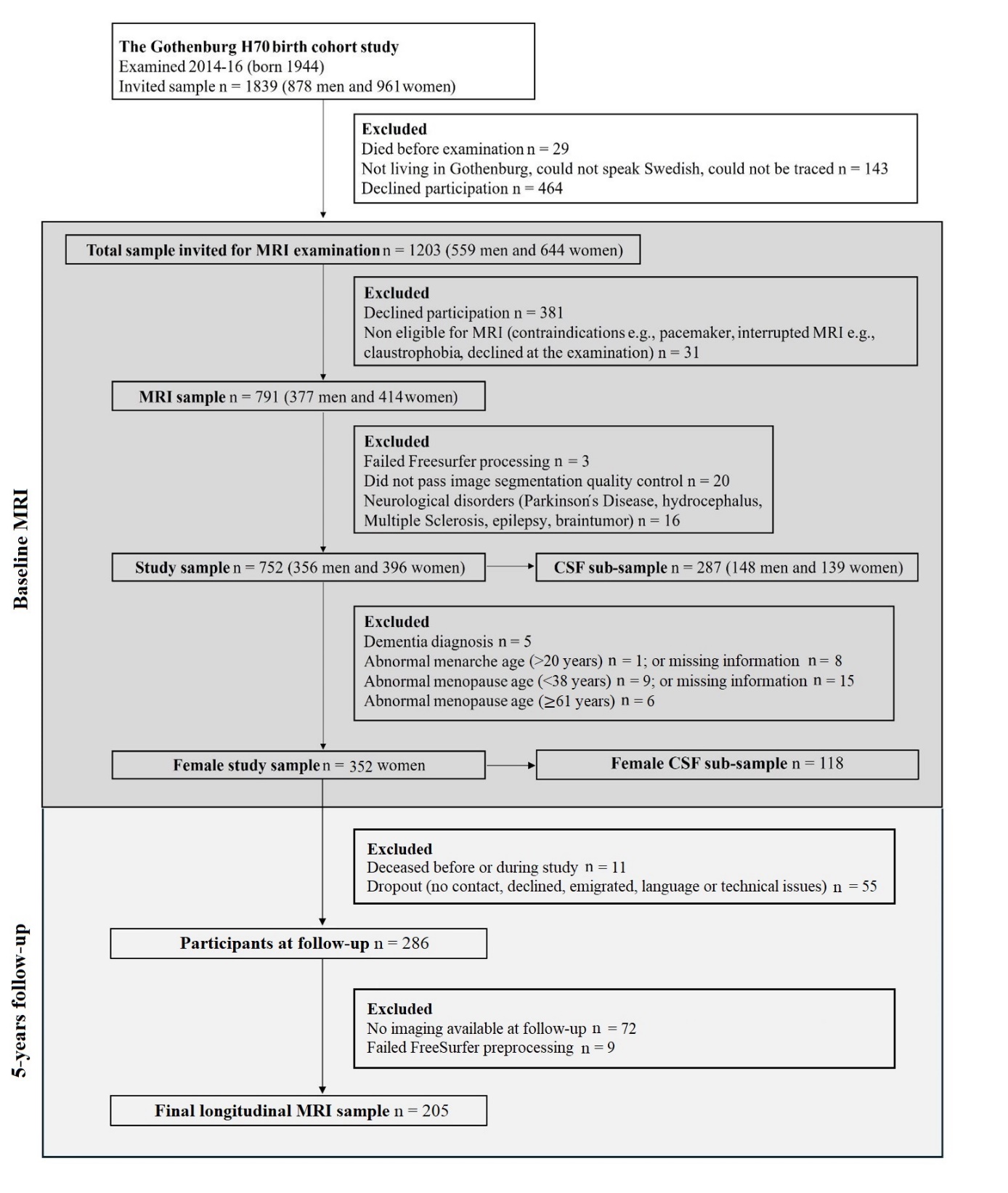

### **Supplementary Figure 2.** Longitudinal trajectories of Alzheimer’s disease thickness signature (A) and white matter hyperintensity volume (B) by hormonal exposures.

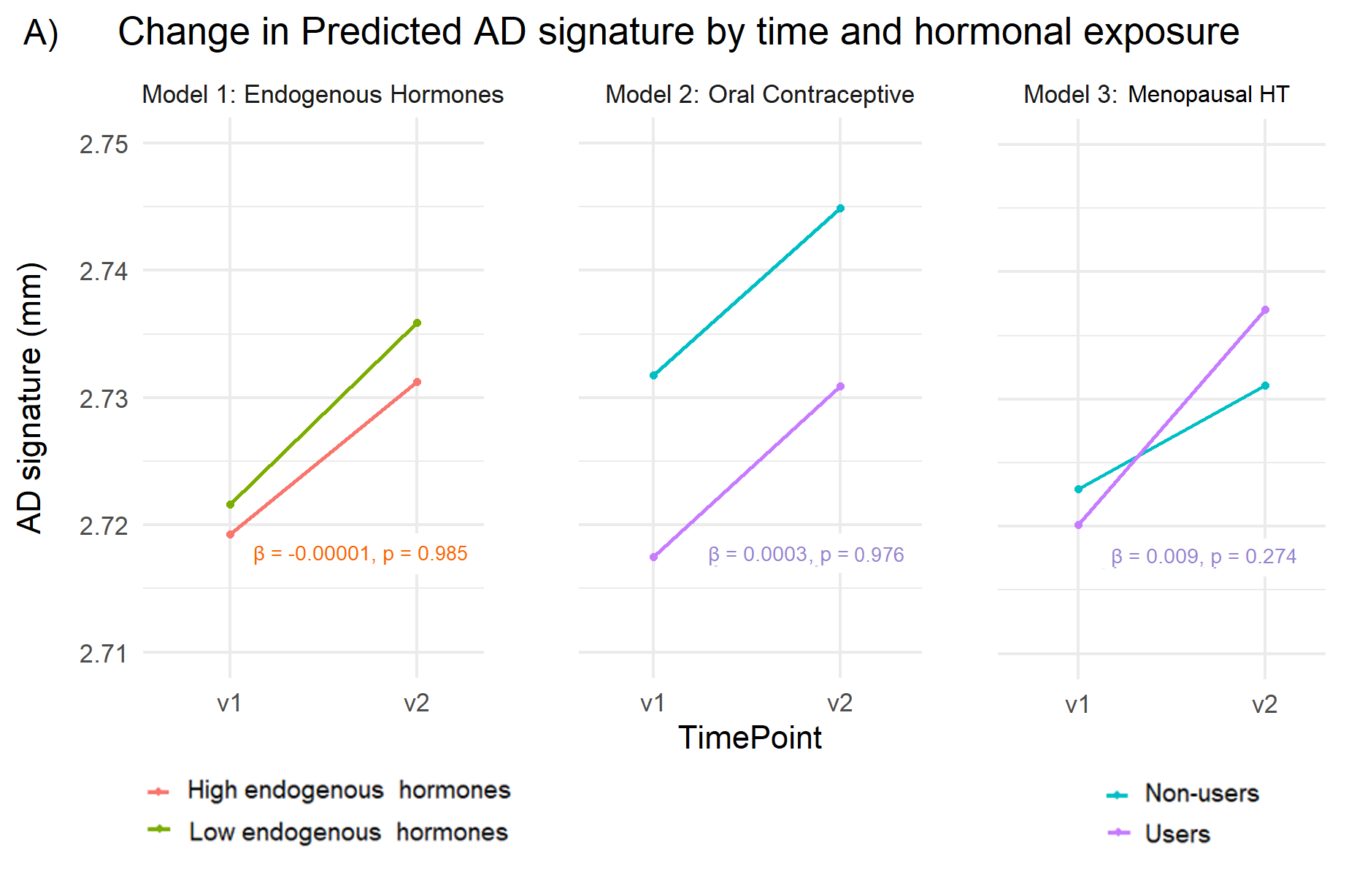

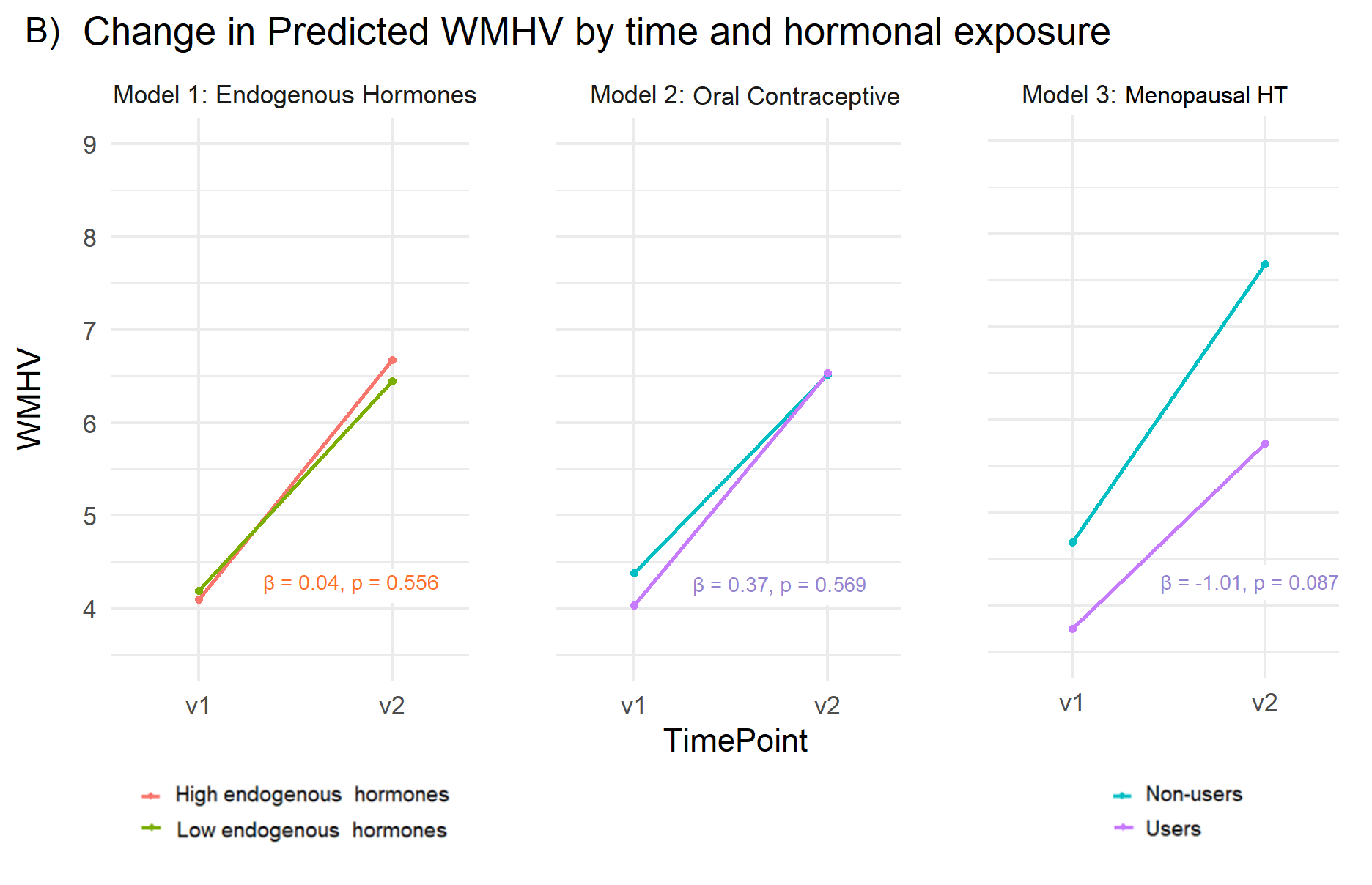
